# Evaluating prescriber preferences for the treatment of indeterminate Chagas disease (EPPIC study): an international survey

**DOI:** 10.64898/2026.09.18.26363397

**Authors:** R Ville-Benavides, C Forsyth, M Certo, S H Gaspar, MJ Pinazo

## Abstract

**Background:** Benznidazole (BNZ) and nifurtimox (NFX) remain the only approved treatments for *Trypanosoma cruzi* infection, but both are associated with frequent adverse drug reactions (ADRs), long treatment duration, and limited accessibility, resulting in poor treatment adherence and premature discontinuation. Recent clinical trials have evaluated shorter and lower-dose regimens; however, the minimum level of efficacy considered acceptable by healthcare providers –in the context of improved safety and greater accessibility –has not been systematically investigated. This study aimed to identify prescriber-defined efficacy thresholds and the factors influencing prescribing preferences for new antiparasitic regimens for Chagas disease.

**Methods:** We conducted an international, cross-sectional, web-based survey between June and August 2024 among healthcare providers, researchers, and public health practitioners with experience in Chagas disease from endemic and non-endemic countries. The survey assessed satisfaction with current antiparasitic drugs, perceived treatment disadvantages, and the willingness of practitioners to prescribe hypothetical new regimens with varying efficacy levels and additional clinical attributes.

**Results:** Thirty of 50 invited participants completed the survey (60% response rate); most were physicians or nurses (57%), and 76% had more than six years of experience managing Chagas disease. An efficacy threshold of 70% or above was associated with substantially higher prescribing willingness: 46.7% of respondents reported being likely or very likely to prescribe a regimen with 70-79% efficacy, compared with 20.0% for 60-69% and 6.7% for 50-59%. Willingness to prescribe regimens with moderate efficacy (60-69%) was increased when regimens offered prevention of mother-to-child transmission, reduced ADR frequency and severity, or the possibility of second-line use. Improved accessibility was an additional determinant for regimen in the lowest efficacy range. Respondents identified ADRs, long treatment duration, and limited drug availability as the main disadvantages of current therapies.

**Conclusions:** This study provides preliminary, provider-level evidence that prescriber acceptance of new antiparasitic regimens for *T. cruzi* infection is shaped by the overall benefit-risk profile, not efficacy alone. A 70% efficacy threshold emerged as a critical inflection point for prescribing willingness, while prevention of mother-to-child transmission, improved tolerability, and greater accessibility were identified as key factors that could support the adoption of regimens with moderate efficacy. These findings may inform the design of future clinical trials and the development of treatment strategies aligned with real-world prescribing priorities.

**Author summary:** Chagas disease affects an estimated 9-11 million people worldwide, yet treatment options remain limited to two approved drugs –benznidazole and nifurtimox –both of which are associated with frequent adverse effects and lengthy treatment courses that are difficult to complete. As new, potentially shorter and better-tolerated regimens are being developed, understanding what level of efficacy healthcare providers consider acceptable is essential for guiding future trial design and treatment development. We surveyed healthcare providers, researchers, and public health practitioners with experience in Chagas disease from both endemic and non-endemic countries. Our findings suggest that a 70% efficacy threshold is critical for prescriber acceptance, and that willingness to prescribe regimens with moderate efficacy can be meaningfully increased when such regimens offer prevention of mother-to-child transmission, reduced adverse effects, or improved accessibility. These insights can inform researchers designing future clinical trials and help align therapeutic strategies with the real-world needs of providers and patients living with *Trypanosoma cruzi* infection.

## Introduction

Chagas disease (CD), caused by the protozoan *Trypanosoma cruzi,* affects an estimated 9-11 million people worldwide and remains a leading cause of infectious cardiomyopathy in Latin America (1, 2). Benznidazole (BNZ) and nifurtimox (NFX) are the only approved treatments for *T. cruzi* infection (3). Although both drugs have demonstrated efficacy in acute and chronic infection (4), their clinical use is limited by a high incidence of adverse drug reactions (ADRs), which contributes to poor adherence and treatment discontinuation (5,6). Between 6% and 31% of patients undergoing the standard 8-week regimen discontinue therapy prematurely, largely due to ADRs (4,7,8).

The pharmacological rationale for exploring shorter or lower-dose BNZ regimens is supported by pharmacokinetic evidence suggesting that the current standard dose may exceed what is necessary for trypanocidal activity (9). The standard 5 mg/kg/day regimen produces mean serum concentrations at the upper limit of the accepted trypanocidal range (3-6 μg/mL), also reported in the same pharmacokinetic study. Altcheh et al. demonstrated higher hepatic clearance and cure rates in children compared with adults, suggesting that equivalent therapeutic responses may be achievable at lower doses in adults (10). A population pharmacokinetic model subsequently confirmed that 2.5 mg/kg/day was sufficient to maintain BNZ concentrations within the therapeutic range (11).

Clinical evidence from recent Phase II trials supports this rationale. In the BENDITA (12) and MULTIBENZ (13) trials, shorter BNZ courses were associated with lower rates of permanent treatment discontinuation compared with the standard 8-week regimen, suggesting a favorable safety profile for abbreviated regimens (12,13). A subsequent re-analysis of data from the BENDITA and E1224 trials showed that the probability of cure was somewhat lower for 2-week regimens than for 4- and 8-week regimens (12,14); however, treatment completion rates were higher in the shorter duration groups, highlighting the inherent trade-off between efficacy and tolerability.

Access to antiparasitic treatment represents an additional and critical barrier: it is estimated that fewer than 1% of people living with *T. cruzi* infection receive treatment (15). This treatment gap reflects not only drug availability constraints but also challenges related to healthcare infrastructure, diagnostic capacity, and the long duration of current regimens.

Taken together, the available pharmacological and clinical evidence suggests that shorter BNZ regimens, or regimens employing reduced daily doses, may offer non-inferior or moderately reduced efficacy while substantially improving tolerability, treatment completion, and ultimately, access. However, the minimum level of efficacy that would be considered acceptable by healthcare providers, in the context of improved safety and greater accessibility, has not been systematically evaluated. This study aimed to assess the minimum acceptable treatment efficacy threshold among experienced healthcare providers in both endemic and non-endemic countries, and to identify the factors that could influence prescribing decisions for new antiparasitic regimens for *T. cruzi* infection.

## Methods

The Evaluating Prescriber Preferences for Indeterminate Chagas treatments (EPPIC) study was an international, cross-sectional, web-based survey conducted between June and August 2024. The survey was administered online via Microsoft Forms and was accessible to participants from both endemic and non-endemic countries.

### Objectives

The primary objective was to estimate the minimum acceptable trypanocidal efficacy of potential new antiparasitic regimens for adults with chronic *T. cruzi* infection in the indeterminate phase, from the perspective of healthcare professionals with experience in CD. Secondary objectives were to: (i) describe respondents’ satisfaction with currently available antiparasitic drugs (BNZ and NFX); (ii) identify the main perceived disadvantages of current treatment options; and (iii) assess factors that could increase willingness to prescribe hypothetical new regimens with lower efficacy.

### Participants’ eligibility criteria

We included adult individuals who self-identified as healthcare providers (e.g., physicians, nurses), researchers, or public health practitioners with experience in CD. Experience was defined broadly to encompass both clinical management and public health roles. Participation was voluntary.

### Recruitment and sampling

We used purposive sampling to ensure representation across professional roles, geographic regions, and gender. Participants were recruited through the professional networks of the study investigators, including DNDi collaborators, treating physicians and nurses from both endemic and non-endemic countries, CD researchers, and members of CD and neglected tropical diseases working groups. A total of 50 individuals were invited to participate. No formal follow-up was conducted with non-responders, and information on reasons for non-participation was not collected.

### Development and content of the survey instrument

We developed a structured questionnaire informed by prior literature on treatment access, adherence, and clinician preferences in CD. The instrument was available in Spanish, English, and Portuguese. Prior to deployment, the questionnaire was piloted among four Chagas-experienced professionals to assess clarity and face validity; the instrument was subsequently shortened and revised for clarity based on their feedback.

The questionnaire covered five domains: (1) sociodemographic and professional profile (e.g., age, country, role, training); (2) CD experience (years in practice, approximate patient volume, drugs used); (3) satisfaction with current treatment options (BNZ and NFX); (4) perceived disadvantages of current therapies; and (5) prescribing preferences for hypothetical new regimens with varying efficacy levels and the influence of additional regimen attributes (e.g., reduced ADR frequency, shorter duration, improved accessibility). The full questionnaire is provided in **S1** Appendix.

### Survey instrument: Key items and scales

Satisfaction with BNZ and NFX was assessed on a 3-point categorical scale (*satisfied*, *not satisfied*, *don’t know*). Perceived efficacy was assessed on a 5-point Likert scale (from *very poor* to *very good*) for each drug, including across adults in general and in women of childbearing potential (WOCBP) in particular. Perceived disadvantages were captured as ranked selections from a predefined list (e.g., ADRs, long treatment duration, limited availability), enabling prioritization across items.

### Data management and statistical analysis

The survey was open from June to August 2024, and the dataset was locked for analysis in October 2024. Survey data were exported from Microsoft Forms into an anonymized dataset; no direct personal identifiers were collected. Data were analyzed using R studio version 4.1.1 (R Foundation for Statistical Computing, Vienna, Austria).

Categorical variables were summarized using frequencies and proportions; age was summarized using the median and interquartile range (IQR).

Given the exploratory nature of the study and the small sample size (N=30), all results are presented descriptively using absolute frequencies and proportions. Age was summarized using the median and interquartile range (IQR). Because survey data was available only as aggregated response frequencies, individual-level and within-participant analysis were not possible.

### Ethical considerations

This study was conducted in accordance with the Declaration of Helsinki. The survey and the methodological approach were sent to the Hospital Clínic Barcelona Ethical Committee (CEim) on 16th August 2022 and, given the study characteristics, the response received on 13th September 2022 was that, due to the study characteristics, a review and approval by an ethical committee was not necessary. In any case, and in accordance with the above-mentioned regulation, access to study data was restricted to the principal investigator and designated collaborators. All participants provided electronic informed consent prior to completing the survey.

## Results

### Participant demographics and professional profile

Thirty individuals completed the survey out of the 50 invited to participate (60% response rate). The median age was 55 years (IQR 39.5-65 years). 40% (n=12) were female. Most participants (80%, n=24) were based in Latin America, and 20% (n=6) in the USA or Europe (Table 1).

**Table 1.** Sociodemographic characteristics.

| **Participant sociodemographic and professional characteristics (N=30)** |  |
| --- | --- |
| Age, median (IQR) | 55 (39.5-65) |
| Gender n (%) |  |
| *Male* | 18 (60) |
| *Female* | 12 (40) |
| Country of origin n (%) |  |
| *Argentina* | 3 (10) |
| *Bolivia* | 5 (17) |
| *Brazil* | 3 (10) |
| *Chile* | 1 (3) |
| *Colombia* | 2 (7) |
| *Guatemala* | 2 (7) |
|  | 6 (20) |
| *Mexico* | 2 (7) |
| *Spain* | 1 (3) |
| *Switzerland* | 1 (3) |
| *United Kingdom* | 2 (7) |
| *United States of America* | 1 (3) |
| *Uruguay* | 1 (3) |
| *Venezuela* |  |
| Highest level of education n (%) |  |
| *College degree* | 3 (10) |
| *Master's degree* | 4 (13) |
| *PhD/doctorate* | 8 (27) |
| *Post-doctorate* | 1(3) |
| *Other/missing* | 14 (47) |
| Current position n (%) |  |
| *Physician* | 16 (53) |
| *Nurse* | 1 (3) |
| *Researcher* | 9 (30) |
| *Public health practitioner* | 3 (10) |
| *Other/missing* | 1 (4) |
| Type of medical specialization, n(%) | **N=16** |
| *Internal medicine* | 2 (12) |
| *Infectious disease* | 5 (31) |
| *Cardiology* | 5 (31) |
| *Family medicine* | 1 (6) |
| *Other/missing* | 3 (20) |

IQR: interquartile range.

Physicians and nurses constituted 57% (17/30) of respondents; the rest were researchers or public health practitioners. We found a high level of training within the sample, with 43% (13/30) of participants holding a postgraduate degree. The most reported specialties were infectious diseases and cardiology 31% (5/30).

### Clinical experience in Chagas disease management

Seventeen participants (57%) reported previous experience directly treating individuals with *T. cruzi* infection. All reported more than one year of treating cases, and 52% (9/17) reported over 10 years of experience managing CD. Regarding patient volume, 47% (8/17) had treated 21-100 cases, and 29% (5/17) had treated more than 100 patients over their careers. BNZ was used by 59% (10/17) of participants, NFX by 29% (5/17) of them, and both drugs by only 12% (2/17), Table 2.

**Table 2.** Respondent experience managing Chagas disease.

| Experience treating Chagas disease (N=17) |  |
| --- | --- |
| Years treating Chagas disease n (%) |  |
| 0-1 | 0 (0) |
| 1-5 | 4 (23) |
| 6-10 | 4 (23) |
| 11 or more | 9 (52) |
| Patients with Chagas disease cared for in a given year n (%) |  |
| 0 | 0 (0) |
| 1-20 | 4 (23) |
| 21-100 | 8 (47) |
| >100 | 5 (29) |
| Drugs used to treat patients with Chagas disease n (%) |  |
| *Benznidazole* | 10 (59) |
| *Nifurtimox* | 5 (29) |
| *Both equally* | 2 (12) |
| *Other* | 0 (0) |
| *None* | 0 (0) |

### Satisfaction and perceived disadvantages of current antiparasitic drugs

Among the 30 respondents, seven (23%; 95% CI 9.9-42.3) reported being satisfied with BNZ and seven (23%; 95% CI 9.9-42.3) with NFX. Dissatisfaction was high for both drugs: 50% (95% CI 31.3-68.7) for BNZ and 57% (95% CI 37.4-74.5) for NFX; the remainder reported uncertainty. The overlapping confidence intervals indicate no meaningful difference in satisfaction between the two drugs **(Figure 1).**

**Figure 1.**
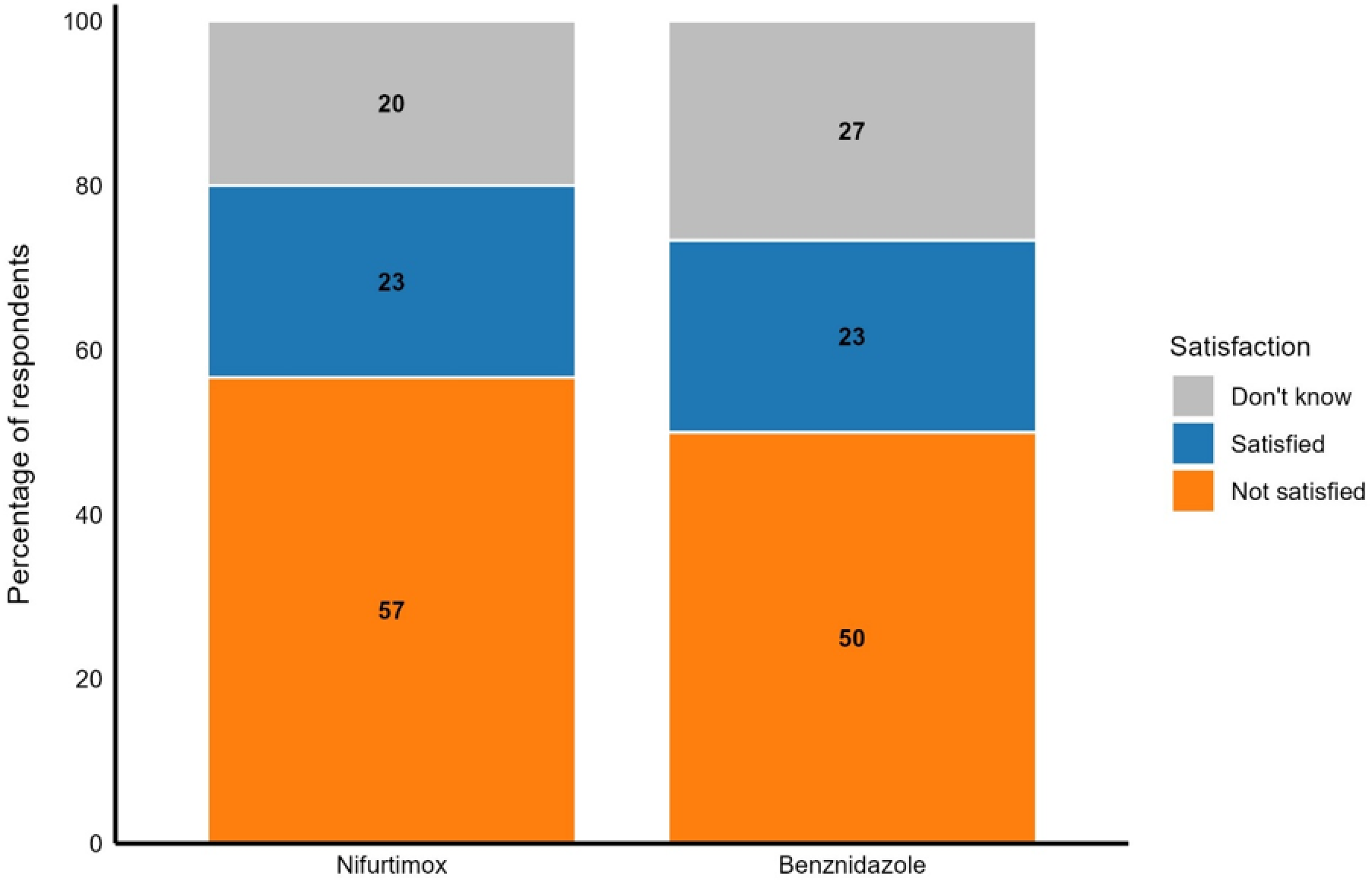
Satisfaction with current antiparasitic treatments for indeterminate Chagas disease. Stacked bar charts show the distribution of responses regarding satisfaction with benznidazole (BNZ) and nifurtimox (NFX) among participants. Response categories were: “Not satisfied”, “Satisfied”, and “Don’t know”. N=30 per treatment group.

The three most cited medication-related disadvantages were (i) the incidence of adverse events, (ii) long treatment duration, and (iii) low availability or difficult access to medicines.

### Perceived efficacy of current treatments

The perceived efficacy of current drugs was rated as “good” for both BNZ and NFX by most respondents; only a minority rated efficacy as “poor” or “very poor.” Perceived efficacy was similar across patient scenarios, including non-pregnant adults and women of childbearing age, suggesting that pregnancy status did not substantially influence respondents’ efficacy perceptions **(Figure 2).**

**Figure 2.**
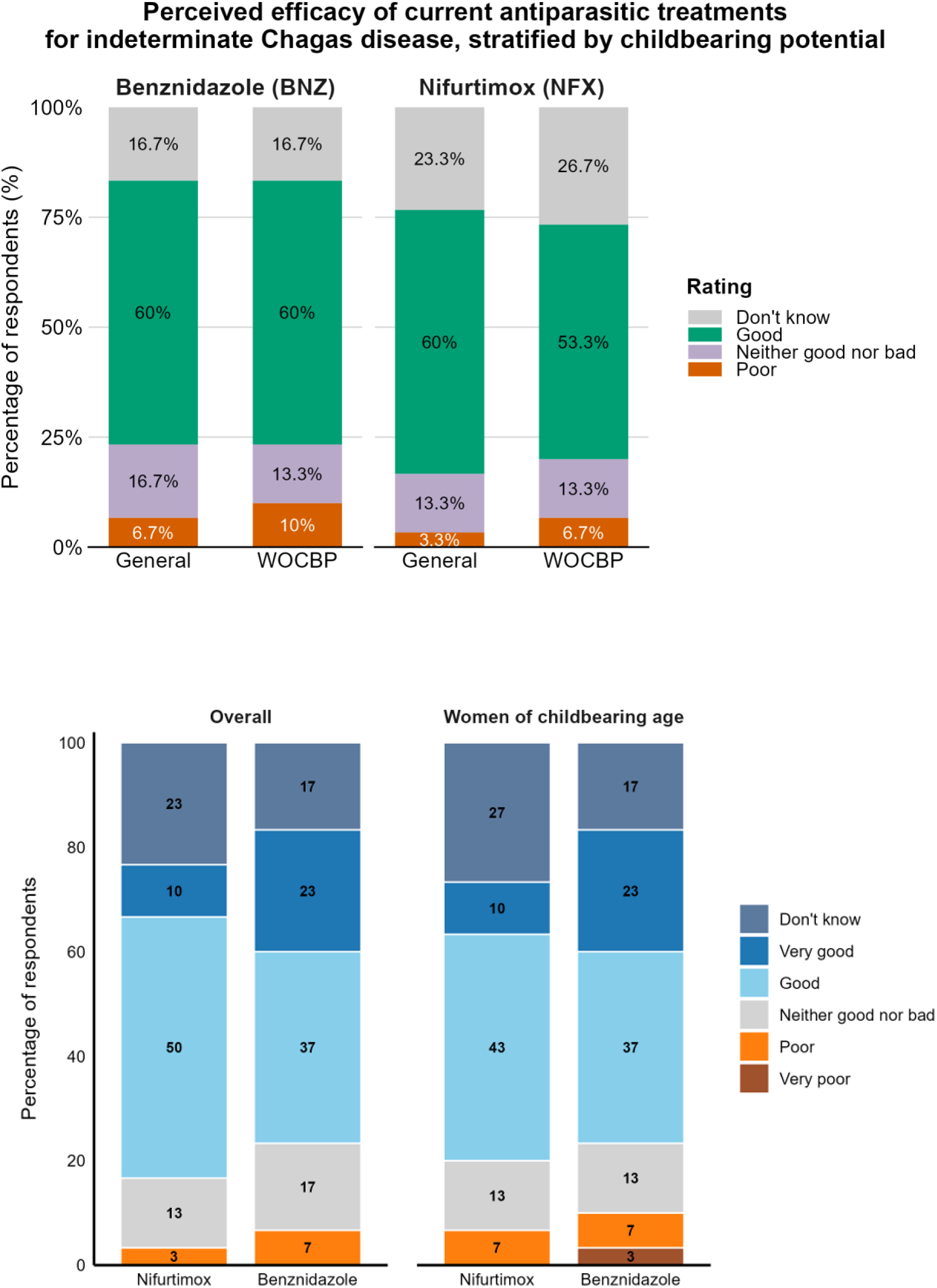
Perceived efficacy of current antiparasitic treatments for indeterminate Chagas disease, by treatment and childbearing potential. Stacked bar charts show the distribution of responses regarding the perceived efficacy of benznidazole (BNZ) and nifurtimox (NFX) among the general population and women of childbearing potential (WOCBP). Response categories were collapsed for clarity: “Poor” combines the original “Very poor” and “Poor” responses; “Good” combines “Good” and “Very good” responses. “Neither good nor bad” and “Don’t know” are presented as originally recorded. N = 30 per treatment group and population.

### Willingness to prescribe new treatments

Participants were asked to rate their willingness to prescribe three hypothetical regimens with decreasing efficacy: Treatment A (70-79%), Treatment B (60-69%), and Treatment C (50-59%). The proportion of respondents reporting a favorable prescribing willingness (”likely” or “very likely”) declined with decreasing efficacy: 46.7% for Treatment A, 20.0% for Treatment B, and 6.7% for Treatment C. **(Figure 3).**

**Figure 3.**
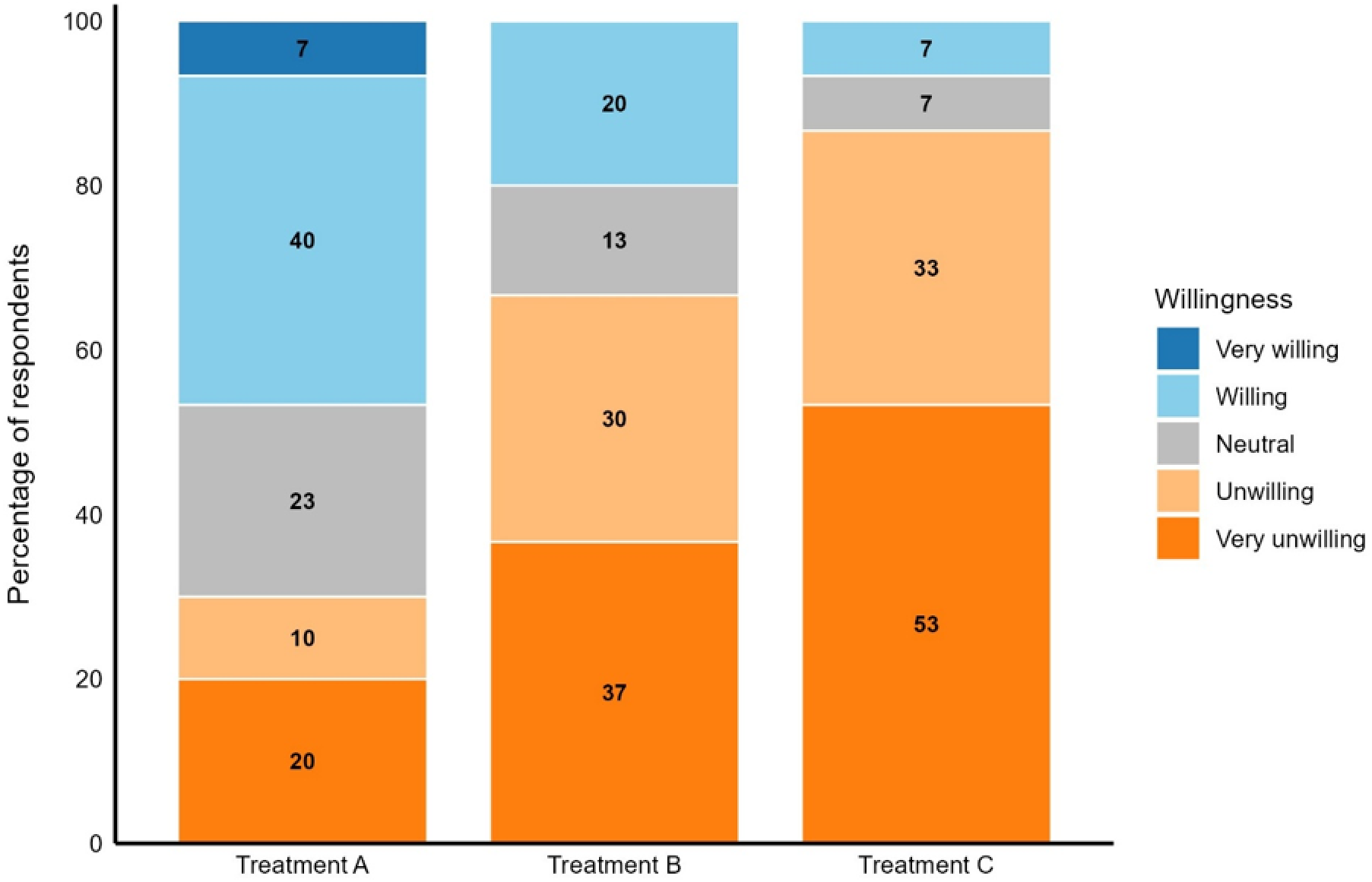
Willingness to prescribe new antiparasitic treatments for indeterminate Chagas disease, by efficacy level. Stacked bar charts show the distribution of responses regarding the willingness of prescribing three hypothetical treatment regimens with decreasing efficacy: Treatment A (70-79%), Treatment B (60-69%), and Treatment C (50-59%). Response options ranged from “Very unlikely” to “Very likely” on a five-point Likert scale. N=30 per treatment group.

When efficacy was below 80%, the top three factors increasing the willingness of prescribing were: prevention of mother-to-child transmission (MTCT), reduced incidence and severity of ADRs, and the possibility of use as second-line therapy. For regimens in the lowest efficacy range (50-59%), greater accessibility emerged as an additional key factor.

Open-text responses indicated that clinicians particularly valued tolerability and treatment accessibility. Several respondents noted that shorter regimens could improve adherence. A few highlighted additional operational challenges, including limited access to PCR-based diagnostic tools and the absence of a reliable test of cure to monitor treatment response.

## Discussion

This cross-sectional survey of healthcare providers, researchers, and public health practitioners with experience in CD revealed areas of concern with existing antiparasitic treatments, including their adverse drug reaction profiles, treatment duration, and accessibility. At the same time, these therapies remain indispensable in current clinical practice and represent the foundation upon which further improvements in treatment can be built.

Perceived efficacy emerged as the dominant factor in prescribing decisions, yet a substantial proportion of respondents indicated willingness to prescribe regimens with moderately reduced efficacy if accompanied by improvements in tolerability, accessibility, or public health impact. These findings provide preliminary, provider-level evidence to inform the design and prioritization of future antiparasitic regimens for *T. cruzi* infection.

The sample demonstrated broad professional and geographic representation, with participation from endemic and non-endemic countries and a range of clinical, research, and public health roles. Over three-quarters of respondents (76%) reported more than six years of experience managing CD, and more than half (52%) reported over ten years of experience, suggesting that the survey captured perspectives informed by substantial clinical expertise.

Regarding the high frequency of ADRs incidence as one of the main concerns in the use of BZD and NFX expressed by professionals that answered the survey, it is important to highlight that in the most of cases, ADRs associated with BNZ and NFX are non-severe and can be managed clinically, allowing people under a treatment regimen to finish the prescribed regimen. It is important to note that mild reactions are often multiple, and may recur after apparent resolution (9), contributing to premature treatment discontinuation in some cases –a pattern consistently observed across clinical trials and observational series (12,16–21). The perceived excessive duration of the standard two-month regimen further compounds adherence challenges. Notably, shorter BNZ courses have been associated with fewer permanent discontinuations in recent Phase II trials, supporting the rationale for abbreviated regimens even if accompanied by modest reductions in efficacy; such regimens are currently being assessed in a Phase III trial (23).

Perceived efficacy was the dominant determinant of prescribing willingness in this survey. The proportion of respondents willing to prescribe declined sharply as hypothetical efficacy decreased: from 46.7% for a regimen with 70-79% efficacy, to 20.0% at 60-69%, and 6.7% at 50-59%. This stepwise pattern suggests that a 70% efficacy threshold represents a critical inflection point below which prescriber acceptance drops substantially. Nevertheless, a meaningful proportion of respondents indicated that their willingness to prescribe a regimen with moderate efficacy (60-69%) could be increased by specific additional attributes –principally the prevention of MTCT, a reduction in ADR frequency and severity, and the possibility of use as a second-line option. For regimens in the lowest efficacy range (50-59%), improved accessibility emerged as an additional modifying factor. These findings suggest that the clinical acceptability of a new regimen is not determined by efficacy alone but by the overall benefit-risk profile as perceived by the prescriber.

Respondents also highlighted important operational barriers to treatment monitoring, including limited access to PCR-based diagnostic tools and the absence of a validated early test of cure. These concerns reflect a broader challenge in CD management: the difficulty of confirming treatment success in routine clinical practice, which further complicates the adoption of regimens with less established efficacy profiles. Future treatment strategies must, therefore, address not only pharmacological performance but also implementation feasibility at the point of care.

These findings are consistent with, and may help to inform, several ongoing research initiatives in the field. An individual participant data meta-analysis being organized by DNDi and the Infectious Diseases Data Observatory (IDDO) aims to refine the evidence base for CD treatment, including the identification of optimal PCR monitoring timepoints and the assessment of geographic variation in treatment response (14). In parallel, clinical and modeling studies are evaluating abbreviated BNZ regimens –including 400 mg/day for two weeks and 300 mg/day for four weeks –through studies such as BenLatino (CuidaChagas) and NuestroBen (DNDi-ELEA) (22,23). The prescriber preference thresholds identified in the present study may provide useful benchmarks for interpreting the clinical relevance of efficacy estimates from these ongoing and future trials.

### Limitations

This study has several limitations that should be considered when interpreting the findings. The sample size was small (N=30), and while purposive sampling enabled representation across professional roles and geographic regions, participants were recruited through investigator networks and may not be representative of the broader CD provider community; selection bias toward providers more engaged with CD research cannot be excluded. Importantly, survey data were available only as aggregated response frequencies exported from the data collection platform, with no access to individual participant-level data. This constrained the statistical analysis to descriptive methods throughout, precluded within-participant testing of prescribing preferences, and limited the ability to explore associations between respondent characteristics and outcomes. Accordingly, all findings should be interpreted as exploratory and hypothesis-generating rather than confirmatory. The greater familiarity of respondents with BNZ compared with NFX –consistent with the predominant use of BNZ in most endemic settings –may mean that satisfaction and limitation ratings for NFX were less precisely informed. Finally, this survey captured the perspectives of healthcare providers only; patient preferences regarding efficacy-tolerability trade-offs may differ and represent an important area for future research. Despite these limitations, the consistency of the observed trends across respondents with diverse backgrounds supports the relevance of these preliminary findings for informing future treatment development.

### Conclusions

In this exploratory, cross-sectional survey of healthcare providers, researchers and public health practitioners experienced in CD, a 70% efficacy threshold emerged as a potential inflection point for prescriber acceptance of new antiparasitic regimens for *T. cruzi* infection, with willingness to prescribe declining below this level. These preliminary findings suggest that the clinical acceptability of new regimens is shaped not by efficacy alone, but by the overall benefit-risk profile –including tolerability, accessibility, and prevention of MTCT. Therapeutic innovation for *T. cruzi* infection should therefore balance trypanocidal efficacy with these broader attributes, and these insights may inform non-inferiority margin selection in future clinical trials and the development of treatment strategies aligned with real-world implementation needs.

## Data Availability

All relevant data are within the manuscript, given that it is a survey and all the data obtained in the responses are all included in manuscript results

## Acknowledgements

The authors would like to sincerely thank all healthcare providers, researchers, and public health practitioners who participated in this survey. We are particularly grateful for the time and expertise shared by individuals across diverse professional roles, geographic regions, and health system contexts. Their perspectives were essential to this study and to advancing understanding of the current landscape of CD management.

## Use of AI tools

We acknowledge the use of Anthropic’s Claude (claude.ai, accessed March 2026) for grammar and English language review, and assistance with code writing and debugging.

## S1 Appendix. Survey about chronic indeterminate Chagas disease treatment efficacy

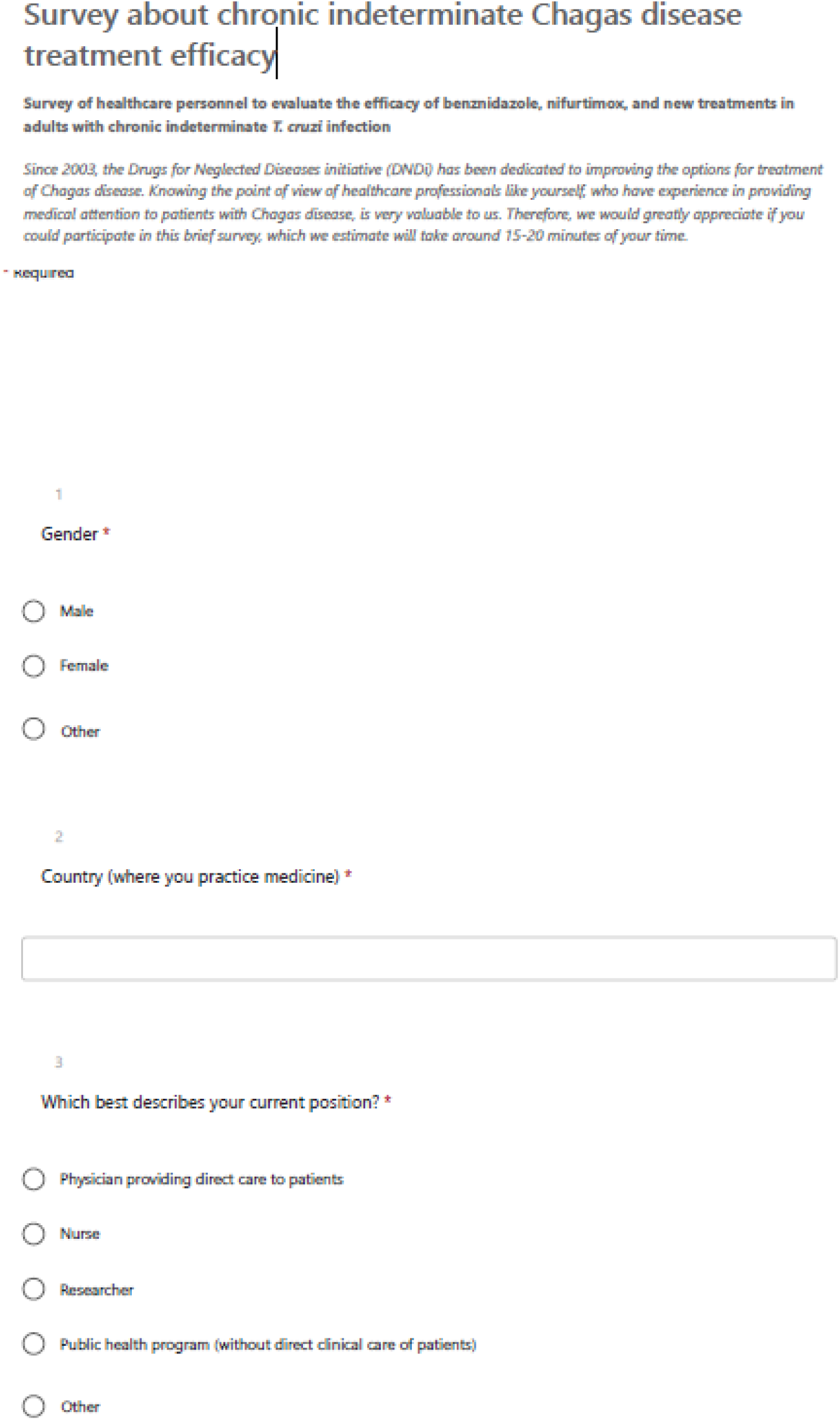
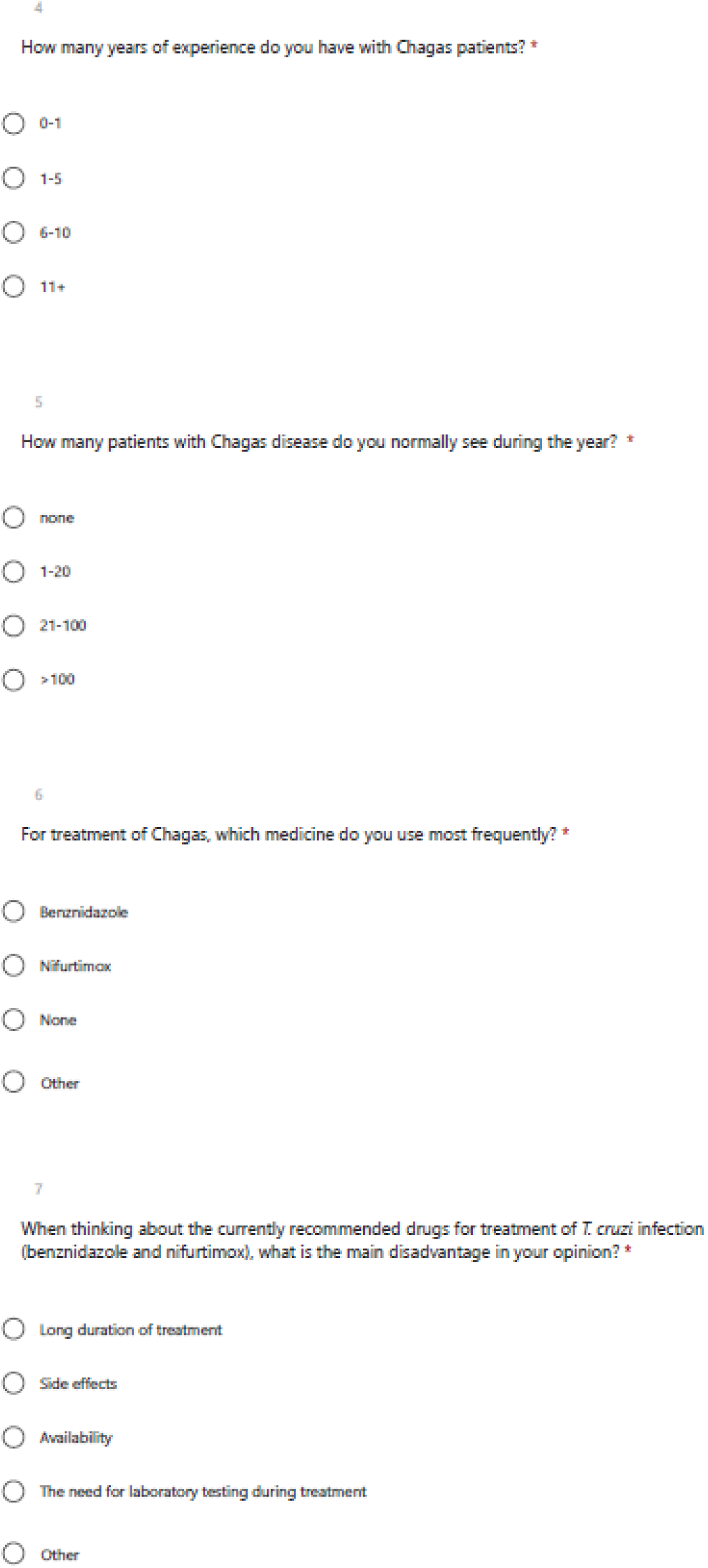
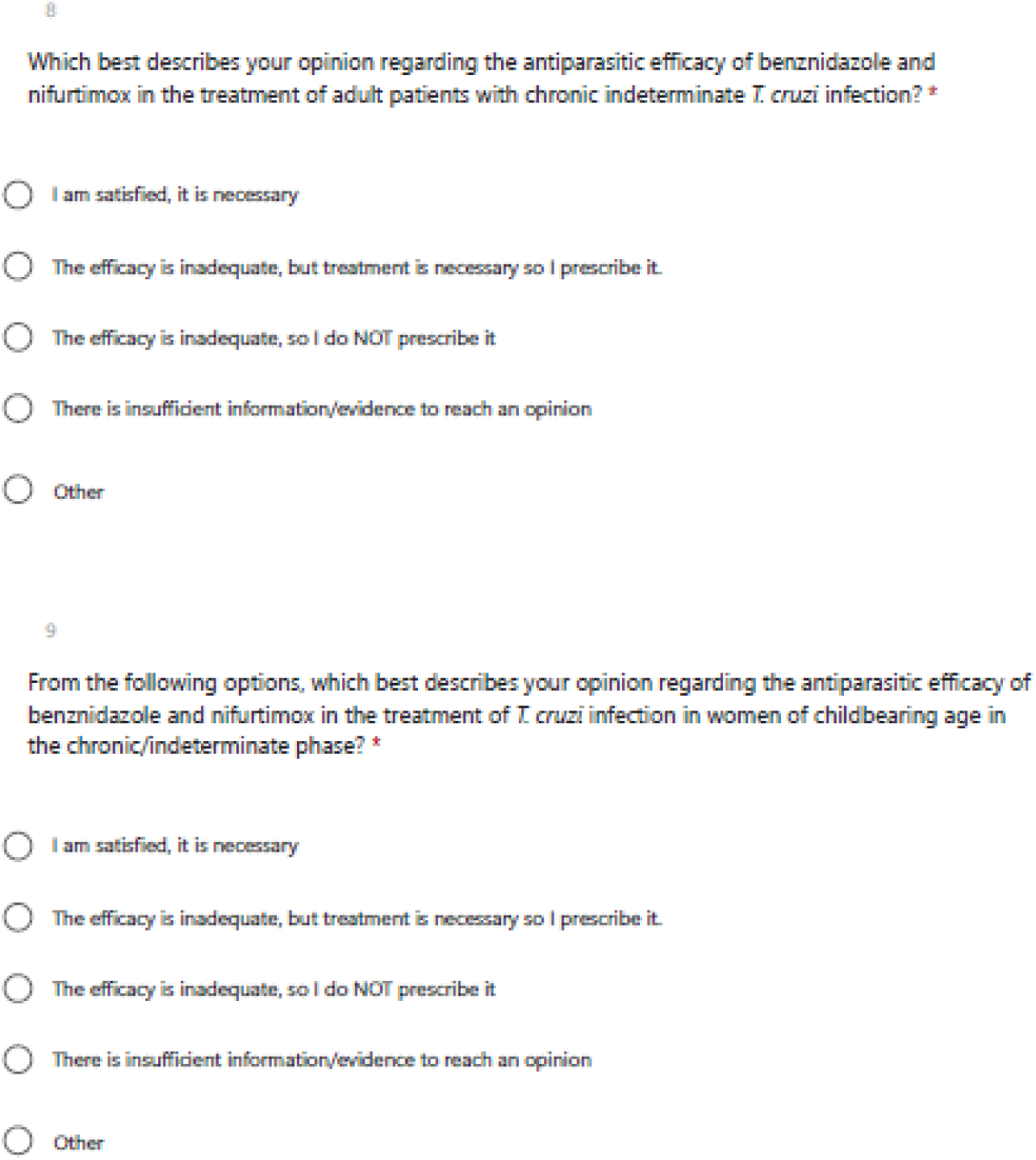
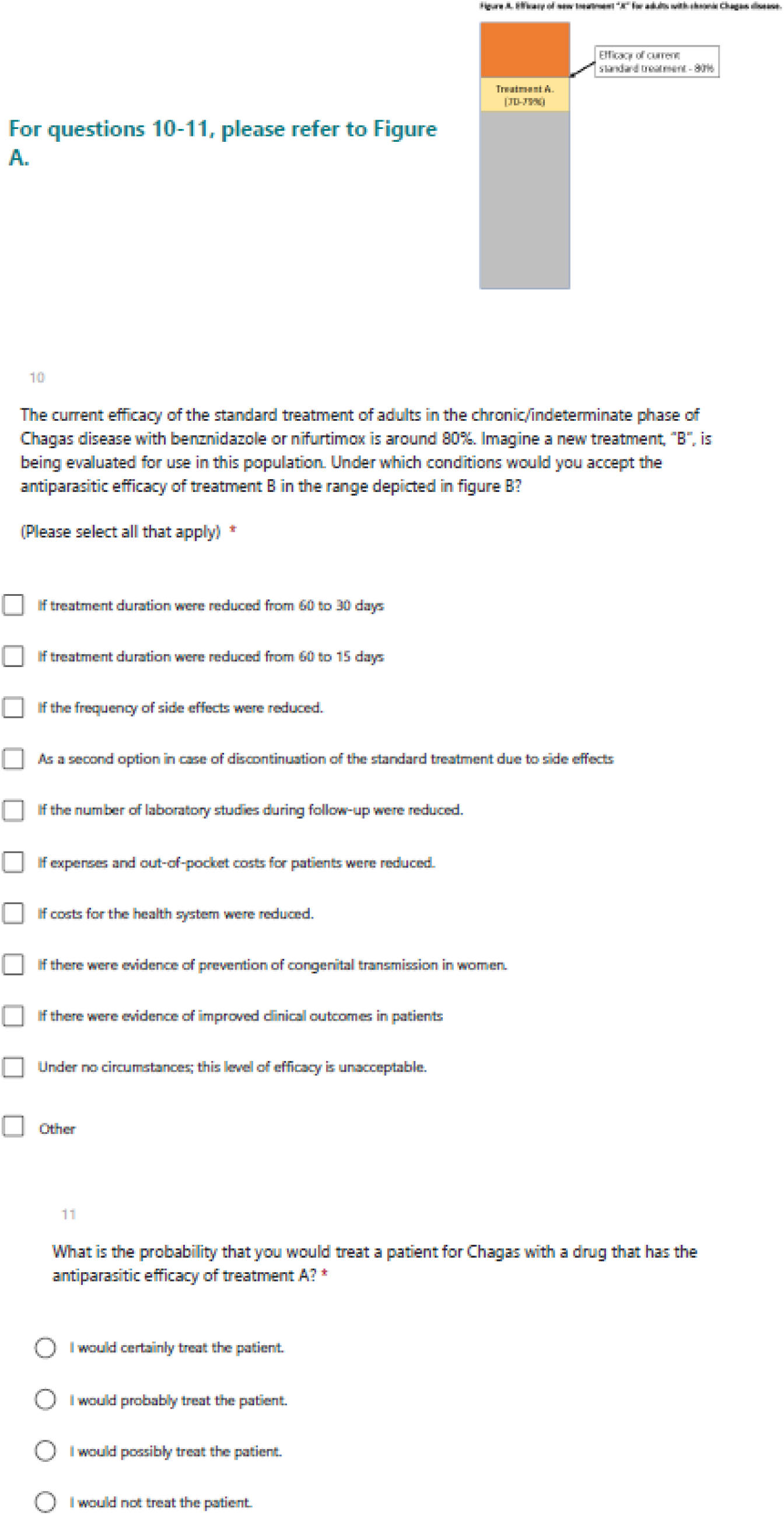
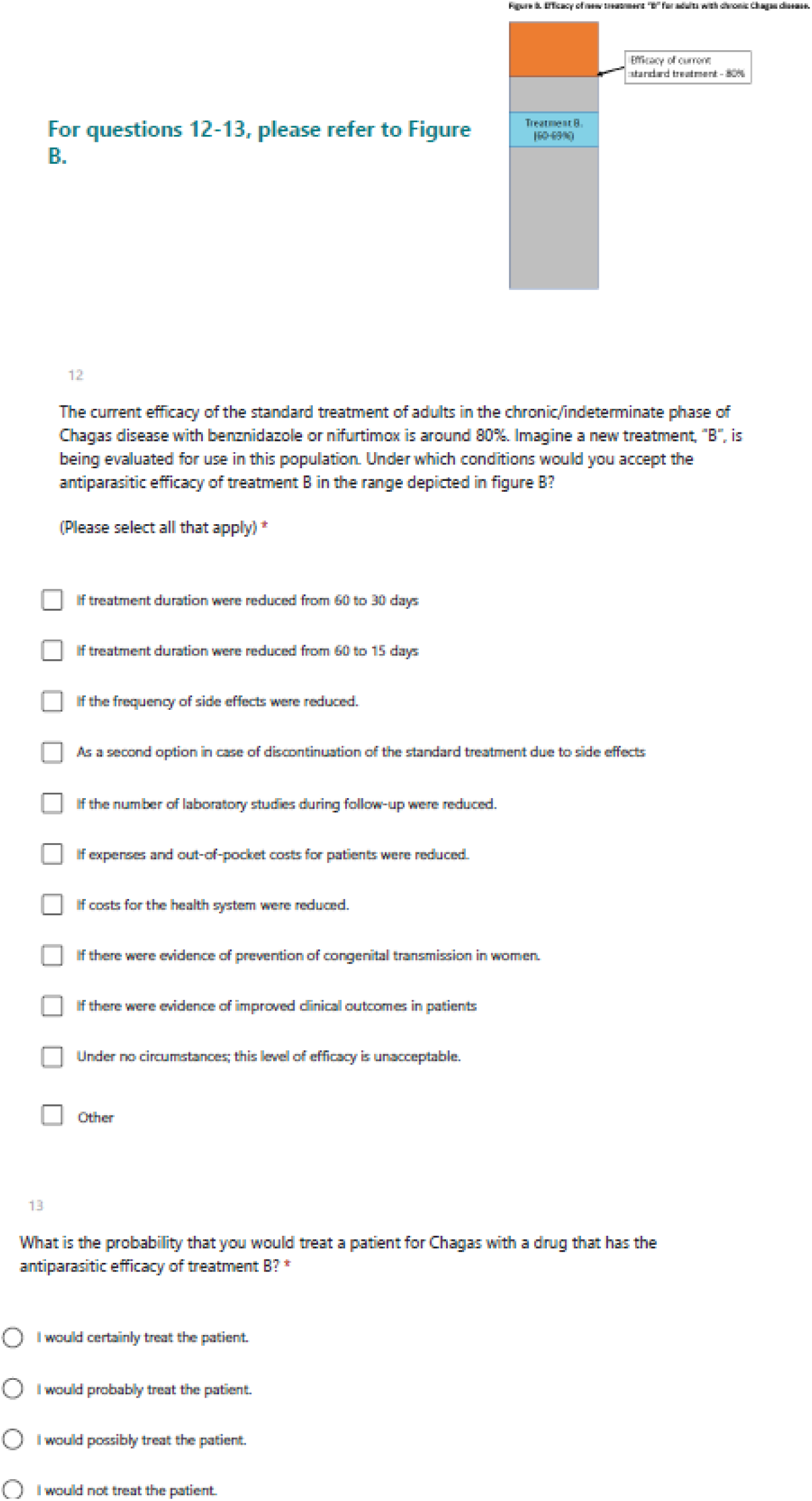
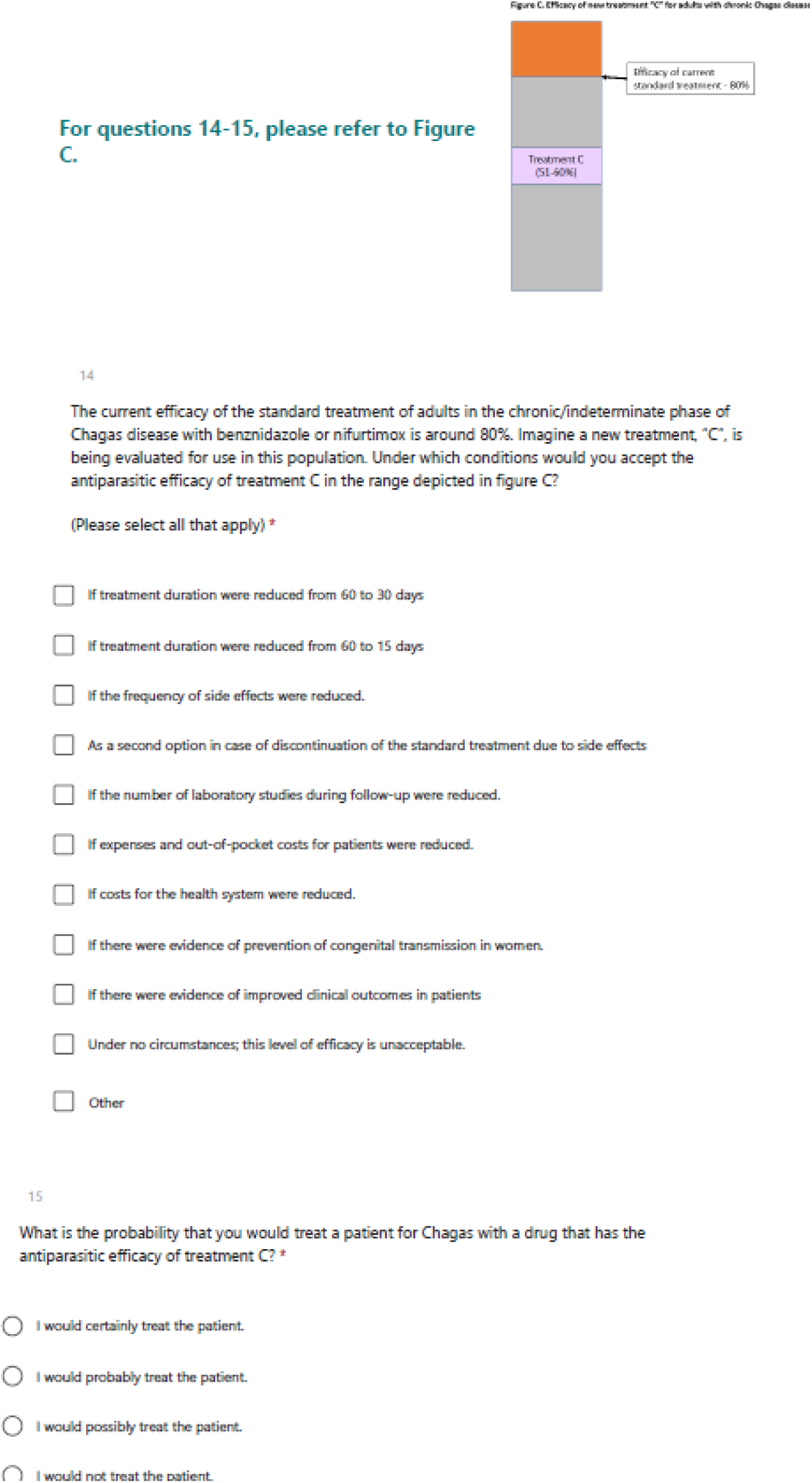
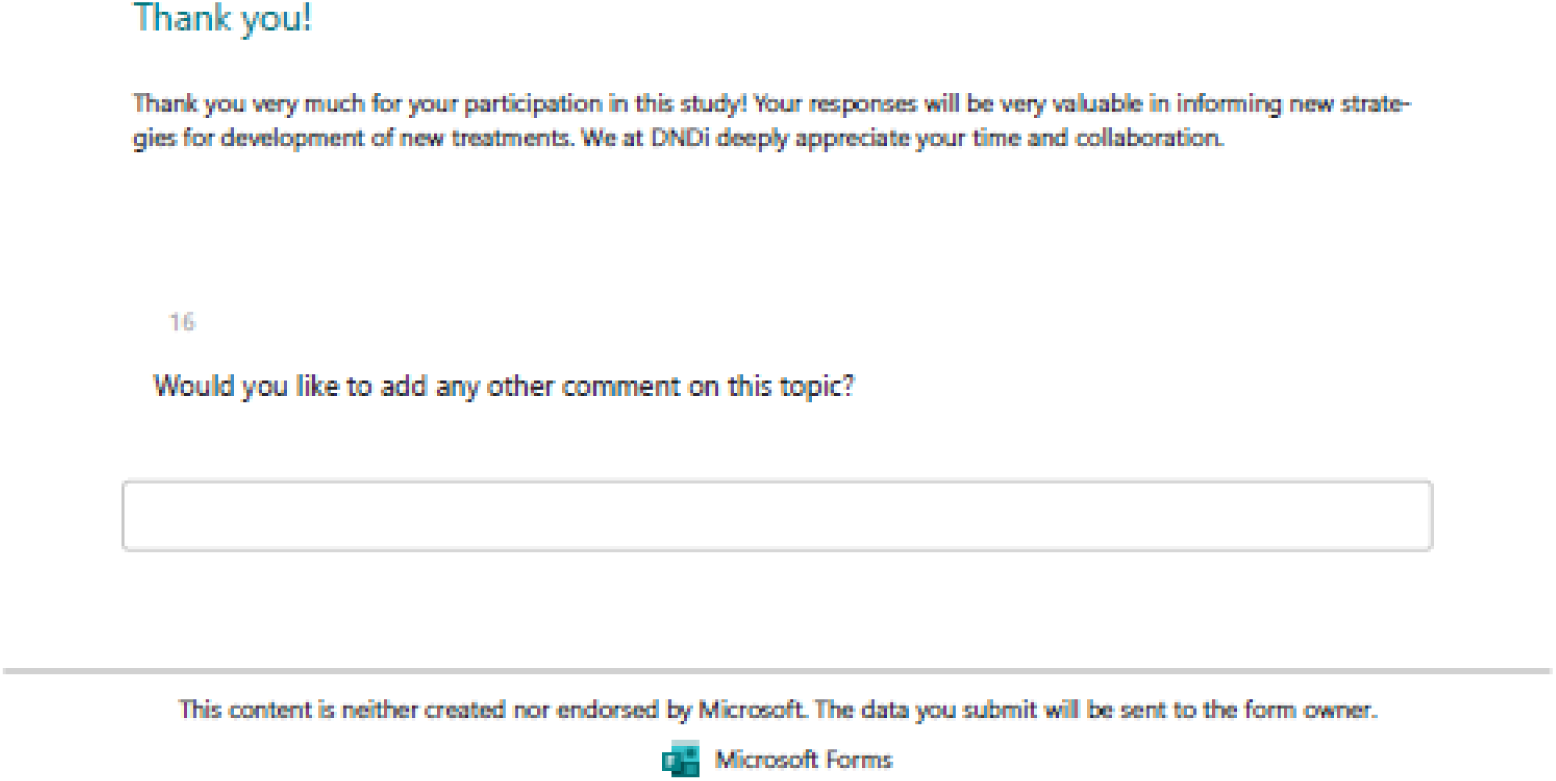

